# Time analysis of dengue deaths that occurred in two regions of Peru during the climatic-atmospheric phenomena El Niño Costero and Cyclone Yaku

**DOI:** 10.1101/2024.03.18.24304491

**Authors:** Willy Ramos, Alfredo Enrique Oyola-García, Aida Aguirre, Jhony A. De la Cruz-Vargas, Miguel Luna, Tania Alarcón, Mónica Meléndez, Juan Huaccho-Rojas, Yudy Cley Cóndor-Rojas

## Abstract

**OBJECTIVE:** To perform a dengue-related death time analysis that occurred in Piura and Ica (Peru) during the atmospheric phenomena of El Niño Costero and Cyclone Yaku in 2023.

**MATERIAL AND METHODS:** A case-series study was performed. We included the deaths between January 1 and June 3, 2023. We reviewed the research data base of the deaths as well as the clinical epidemiology records, death certificates and laboratory test results. A time analysis was performed from the date of onset of symptoms, date of first visit at a health establishment, date that dengue diagnosis was first registered and date of death.

**RESULTS:** We included 40 deaths by dengue in the study. 70% were females, the median age of death was 51 years of age, and 70% came from Piura. 51.2% presented some comorbidity and 14% had self-medicated with metamizole, other NSAIDS or corticoids before the initial medical consultation. 37.5% presented an unconventional clinical pattern at admission and dengue was not suspected in 40% during the initial assessment. The median time until the initial assessment, the diagnosis time, and the time from diagnosis until death were 3 days, 0 days, and 1 day, respectively. Adults presented a time until the initial assessment significantly greater than the other age groups (p=0.048), the diagnosis time was significantly less than those seen at the Health Department establishments and regional government establishments (MINSA/GORE) than in private establishments (p=0.014) and greater in pediatrics (p=0.018). The time from diagnosis until death was significantly less in those that self-medicated before the initial assessment (p=0.041).

**CONCLUSION:** The adults presented a significantly greater time until the initial assessment; the diagnosis time was less than those seen in MINSA/GORE establishments and greater in pediatrics, furthermore, the time from diagnosis until death was significantly less in those who self-medicated before the initial assessment. An important fraction of deaths presented comorbidities and an unconventional clinical pattern at admission, with dengue frequently not suspected during the initial assessment.

## INTRODUCTION

Dengue constitutes a public health problem of great magnitude in Peru, mainly in the regions of the northern Coas, central Coast, and Amazon. The existing favorable weather conditions at the end of spring and summer in these regions, the impact of social determinants such as poverty, overcrowding, and lack of drinking water, as well as the existing political instability in the last years difficult its prevention and control.^1-4^

During the first half of 2023, Peru faced the effects of the “El Niño Costero” phenomenon (FENC), which was characterized by a concentrated increase in the sea surface temperature in the north and central coast, an increase in atmospheric temperature, and torrential rains, which caused massive flooding and overflowing of rivers, mainly in the country’s northern ^1,5,6^. On the other hand, the Cyclone Yaku was an atmospheric phenomenon that affected the northern and central coast of Peru since the last week of February until the third week of March 2023, that increased the rain caused by FENC which brought flooding and overflowing of rivers as a consequence ^7^. In this context, the Peruvian government issued a state of emergency to mitigate the consequences of disasters, which included reorganizing health services^8,9^.

The increase of environmental temperature, as well as the existence of flood zones favored the reproduction of the *Aedes aegypti* vector, which caused the dengue epidemic of greatest magnitude ever registered in Peru, greater than the one during FENC of 2017 ^10-13^. In a similar manner as what occurred during the COVID-19 pandemic, the great number of dengue cases overburdened the health care facilities in the northern coast of the country, mainly the departments with a lower response capacity ^6,7,14^.

During epidemiological (EPI) week of 2023, 59,145 confirmed dengue cases without warning signs, 7,329 with warning signs, and 280 severe dengue cases were reported, likewise, 58,771 probable cases without warning signs, 5,147 with warning signs, and 154 severe dengue cases were reported. For this EPI week, 201 deaths had accumulated during the year, which was triple of that notified up to EPI week 22 2022, in which 60 deaths occurred due to dengue. It was also greater than the total number of deaths due to dengue during the year 2017 (89 deaths) in which one FENC had occurred ^10,15^. According to the situation room at the National Center of Epidemiology, Prevention and Control of Diseases of the Health Department of Peru (CDC-Peru), EPI week 21 2023, the regions with greater number of deaths by dengue were Piura (49 deaths), Lambayeque (34 deaths), and Ica (33 deaths).

The objective of this study was to analyze the times of deaths by dengue occurring in regions of Piura and Ica during El Niño Costero and Cyclone Yaku climatic-atmospheric phenomena that contributed to understanding the reasons of high mortality by dengue in Peru during the year 2023.

## MATERIAL AND METHODS

A case series study was performed. We included the deaths by dengue from EPI week 01 until EPI week 22 (between January 1^st^ and June 3^rd^, 2023) in Ministry of Health/Regional Government (MINSA/GORE) establishments and private establishments from the regions of Piura and Ica, which were among the most affected by El Niño Costero and Cyclone Yaku climatic phenomena.

We included in the study the deaths that fulfilled the following criteria:

- Deaths occurred as a consequence of the clinical signs or the clinical evolution of dengue during the medical care process, or in how dengue significantly contributed to the clinical worsening of another disease.
- Dengue confirmation through laboratory tests (RT-PCR, ELISA IgM/IgG) or an epidemiological nexus.
- Deaths that did not have laboratory test confirmation or an epidemiological nexus reviewed by a medical team for its classifications, which concluded that the clinical signs corresponded to dengue.

We excluded deaths that presented the following criteria:

- Cases in which death was explained by another disease or in that dengue did not influence its clinical course.
- Deaths with insufficient clinical information to attribute dengue as the direct or indirect cause of death, even if there were positive results in laboratory tests and/or in an epidemiological nexus.

The sample was comprised of 44 deaths by dengue obtained from the database of clinical-epidemiological research sheets from the dengue epidemiological surveillance (31 from Piura and 13 from Ica). This database was obtained by consecutive sampling from the population of 69 deaths by dengue (45 from Piura and 24 from Ica).

We reviewed the clinical-epidemiological research data of deaths as well as the information from clinical-epidemiological research surveillance sheets, death certificates and laboratory test results. We obtained epidemiological, clinical, and medical care variables from this database:

- Epidemiological variables: Age, sex, place of residence, gestation, diagnosis of comorbidities, self-medication, health provider.
- Clinical variables: length of disease, clinical pattern of presentation, warning signs and severity at admission, thrombocytopenia, hemoconcentration, medical treatment with metamizole or other NSAIDs during medical care.
- Medical care variables: Type of admission, time until blood count was requested, dengue suspected during initial assessment, admission into Intensive Care Unit (ICU).

A time analysis was carried out from the onset of symptoms, date of initial assessment at a healthcare establishment, date in which the medical report reflects the dengue diagnosis (presumptive or definitive) for the first time, and the date of death:

- Time to initial assessment: Time transpired from the onset of symptoms until the initial assessment in a healthcare establishment. We took into account the delay until the initial assessment if this occurred at least 5 days after the onset of symptoms.
- Time of diagnosis: Elapsed time from the initial assessment until it was reported as a dengue diagnosis for the first time.
- Time of diagnosis-death: Elapsed time from the first time it was reported as a dengue diagnosis until the death of the case.

The statistical analysis was performed with the SPSS 26 program for Windows. A univariate statistic was performed based on the frequencies, percentages, median, interquartile range (IQR) obtained, and a bivariate statistic with the non-parametric Mann-Whitney U and Kruskal Wallis tests. A multivariate statistic was performed with a negative binomial regression model with a robust estimator where the dependent variables were time and independent variables were the possible predictors of time obtaining beta coefficients and confidence intervals. The calculations were carried out with a confidence level of 95%.

Since the research was carried out from secondary sources, it did not imply important risks. We guaranteed the confidentiality of the information obtained which was used for the purpose of this study respecting the ethical principles as stipulated in the Declaration of Helsinki. This study was approved by the Research Ethical Committee of the Ricardo Palma University Medical School (Expedited review: PI-010-2023).

## RESULTS

We reviewed 43 deaths from confirmed or probable dengue cases, of which 2 were excluded due to other causes explaining cause of death, and 1 in which it was not possible to establish if death was related to dengue. Given this, 40 deaths from dengue remained available for analysis.

Of the deaths researched, 70.0% were of feminine sex observing great variability in age of death from 2 years of age up to senior adults of 92 years of age (median 51 years), 70.0% resided in the Piura region. Regarding personal history, 51.2% presented comorbidity, mainly hypertension (18.6%), diabetes mellitus (16.0%), chronic kidney disease (11.6%), and hepatic cirrhosis (9.2%). Also, two of the deaths were pregnant women. 14.0% of deaths had self-medicated with metamizole, other NSAIDs or corticoids prior to the initial assessment (Table 1).

**TABLE 1:**
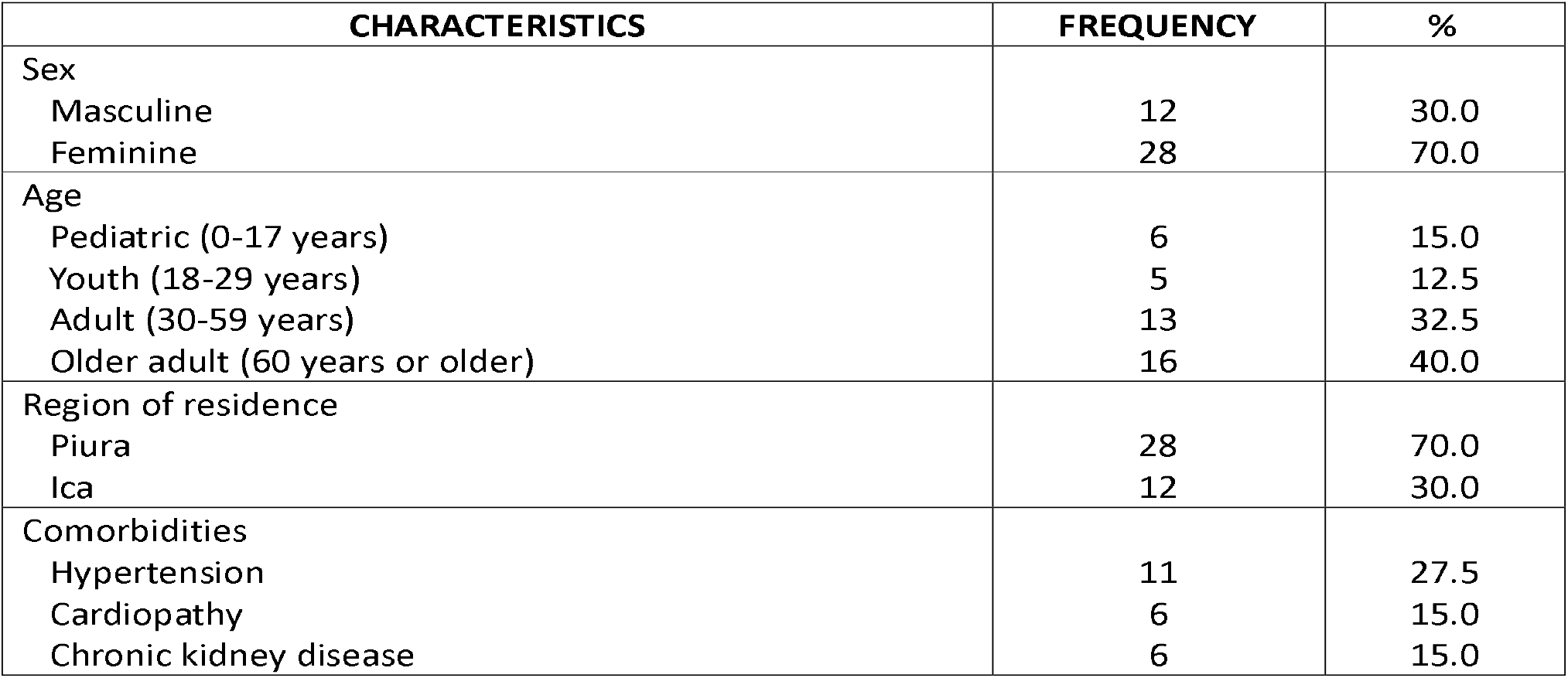
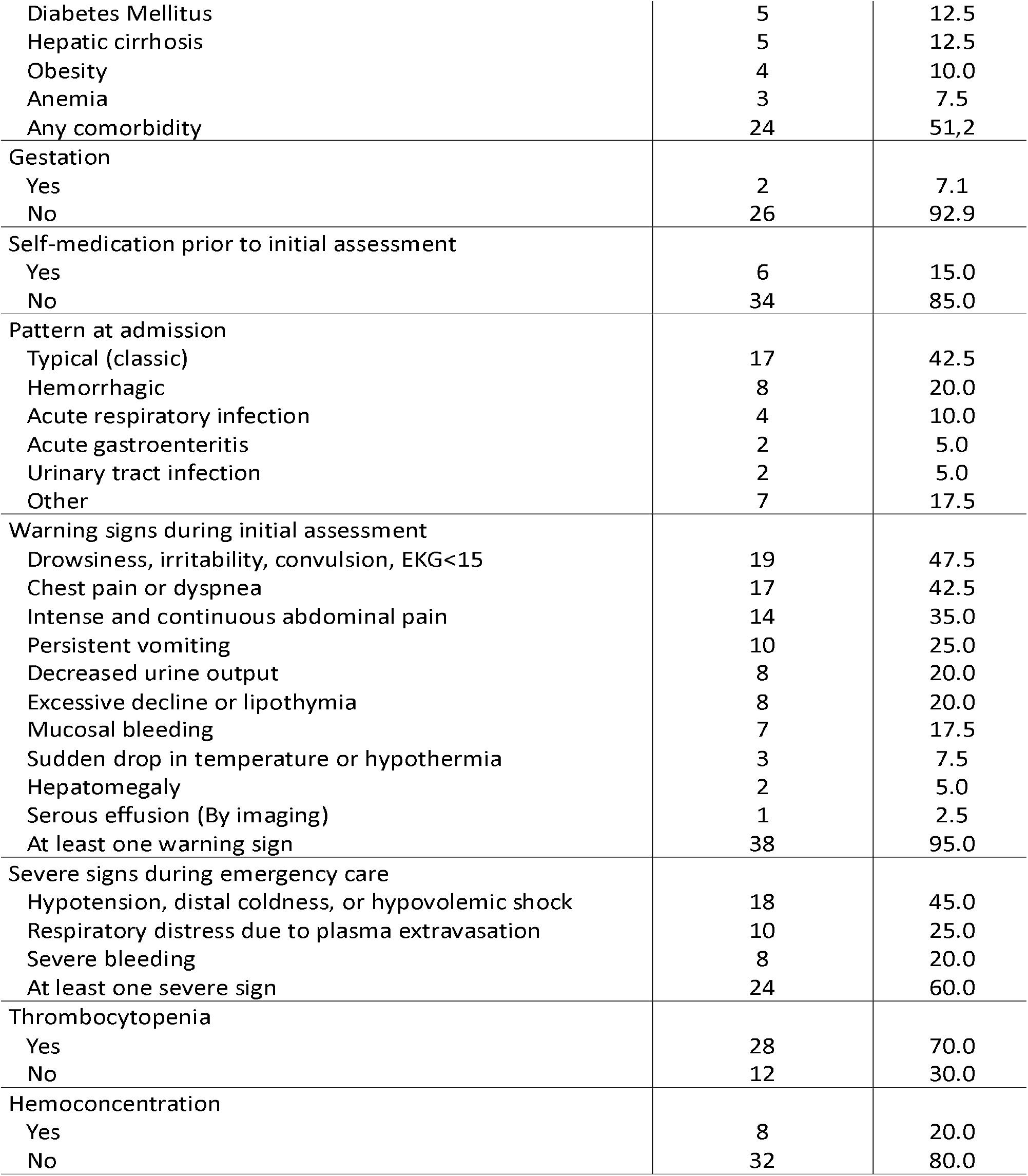
Epidemiological and clinical characteristics of the deceased from dengue in the departments of Piura and Ica.

| CHARACTERISTICS | FREQUENCY | % |
| --- | --- | --- |
| Sex |  |  |
| Masculine | 12 | 30.0 |
| Feminine | 28 | 70.0 |
| Age |  |  |
| Pediatric (0-17 years) | 6 | 15.0 |
| Youth (18-29 years) | 5 | 12.5 |
| Adult (30-59 years) | 13 | 32.5 |
| Older adult (60 years or older) | 16 | 40.0 |
| Region of residence |  |  |
| Piura | 28 | 70.0 |
| Ica | 12 | 30.0 |
| Comorbidities |  |  |
| Hypertension | 11 | 27.5 |
| Cardiopathy | 6 | 15.0 |
| Chronic kidney disease | 6 | 15.0 |
| Diabetes Mellitus | 5 | 12.5 |
| Hepatic cirrhosis | 5 | 12.5 |
| Obesity | 4 | 10.0 |
| Anemia | 3 | 7.5 |
| Any comorbidity | 24 | 51.2 |
| Gestation |  |  |
| Yes | 2 | 7.1 |
| No | 26 | 92.9 |
| Self-medication prior to initial assessment |  |  |
| Yes | 6 | 15.0 |
| No | 34 | 85.0 |
| Pattern at admission |  |  |
| Typical (classic) | 17 | 42.5 |
| Hemorrhagic | 8 | 20.0 |
| Acute respiratory infection | 4 | 10.0 |
| Acute gastroenteritis | 2 | 5.0 |
| Urinary tract infection | 2 | 5.0 |
| Other | 7 | 17.5 |
| Warning signs during initial assessment |  |  |
| Drowsiness, irritability, convulsion, EKG<15 | 19 | 47.5 |
| Chest pain or dyspnea | 17 | 42.5 |
| Intense and continuous abdominal pain | 14 | 35.0 |
| Persistent vomiting | 10 | 25.0 |
| Decreased urine output | 8 | 20.0 |
| Excessive decline or lipothymia | 8 | 20.0 |
| Mucosal bleeding | 7 | 17.5 |
| Sudden drop in temperature or hypothermia | 3 | 7.5 |
| Hepatomegaly | 2 | 5.0 |
| Serous effusion (By imaging) | 1 | 2.5 |
| At least one warning sign | 38 | 95.0 |
| Severe signs during emergency care |  |  |
| Hypotension, distal coldness, or hypovolemic shock | 18 | 45.0 |
| Respiratory distress due to plasma extravasation | 10 | 25.0 |
| Severe bleeding | 8 | 20.0 |
| At least one severe sign | 24 | 60.0 |
| Thrombocytopenia |  |  |
| Yes | 28 | 70.0 |
| No | 12 | 30.0 |
| Hemoconcentration |  |  |
| Yes | 8 | 20.0 |
| No | 32 | 80.0 |

Regarding clinical deaths, 37.5% presented a clinical pattern at admission different from the classic or hemorrhagic and 95.0% presented at least a warning sign. 60.0% presented severe signs during emergency care, 70.0% presented thrombocytopenia in the first blood count and 20.0% presented hemoconcentration.

With respect to the care characteristics, the majority of deceased were seen in hospitals (57.5%), in MINSA/GORE public establishments (82.5%), mainly admitted through emergency (67.5%). In 40.0% of deaths, dengue was not suspected in initial assessment with an elapsed time between admission and the presumptive or definitive diagnosis. In 92.5% of deaths, a blood count was performed within the first 12 hours of admission, 17.5% received treatment for fever with metamizole, and 32.5% were admitted to an ICU. This is shown in Table 2.

**TABLE 2:**
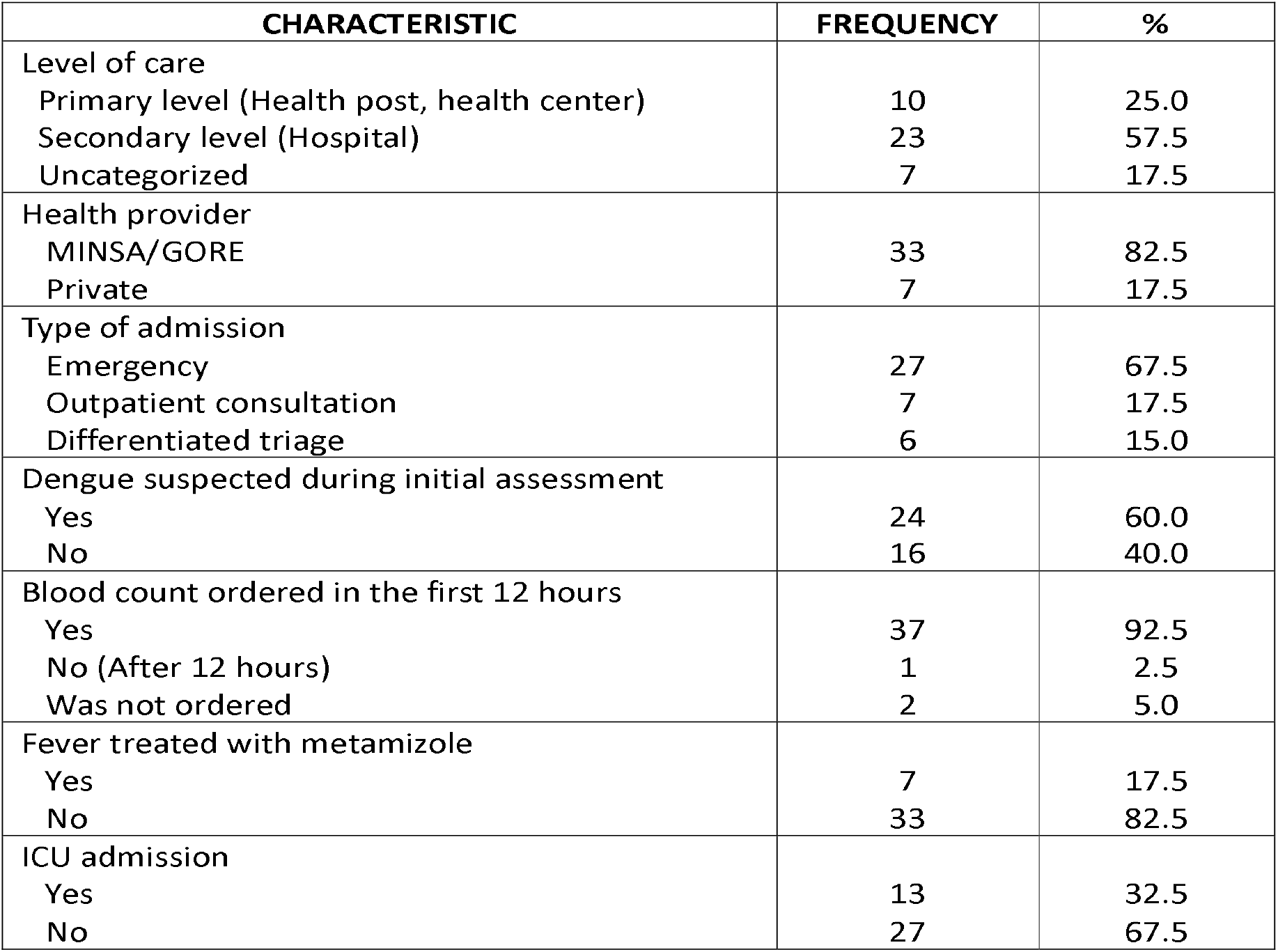
Characteristics of emergency care of those deceased from dengue in the departments of Piura and Ica.

| CHARACTERISTIC | FREQUENCY | % |
| --- | --- | --- |
| Level of care |  |  |
| Primary level (Health post, health center) | 10 | 25.0 |
| Secondary level (Hospital) | 23 | 57.5 |
| Uncategorized | 7 | 17.5 |
| Health provider |  |  |
| MINSA/GORE | 33 | 82.5 |
| Private | 7 | 17.5 |
| Type of admission |  |  |
| Emergency | 27 | 67.5 |
| Outpatient consultation | 7 | 17.5 |
| Differentiated triage | 6 | 15.0 |
| Dengue suspected during initial assessment |  |  |
| Yes | 24 | 60.0 |
| No | 16 | 40.0 |
| Blood count ordered in the first 12 hours |  |  |
| Yes | 37 | 92.5 |
| No (After 12 hours) | 1 | 2.5 |
| Was not ordered | 2 | 5.0 |
| Fever treated with metamizole |  |  |
| Yes | 7 | 17.5 |
| No | 33 | 82.5 |
| ICU admission |  |  |
| Yes | 13 | 32.5 |
| No | 27 | 67.5 |

In the time analysis, we found the longest time was up to the initial assessment that had a median of 3 days, likewise, we observed that 20.0% presented a delay in going to the health establishments to receive the initial care for dengue.

The diagnosis time had a median of 0 days which indicates that the time from admission to the health establishment until the presumptive or definitive diagnosis of dengue was reflected in the medical report was less than 1 day. Likewise, the diagnosis-death time was very short with a median of 1 day (See table 3).

**TABLE 3:** Analysis of times of deaths by dengue in the departments of Piura and Ica.

| TIME EVALUATED<br>(DAYS) | MEDIAN<br>(RIQ) | MINIMUM<br>VALUE | MAXIMUM<br>VALUE |
| --- | --- | --- | --- |
| Time until initial assessment | 3 (1 – 4) | 0 | 8 |
| Time of diagnosis | 0 (0 – 1.8) | 0 | 6 |
| Time of diagnosis-death | 1 (0 – 2) | 0 | 20 |
| Total time | 6 (4 – 7) | 0 | 21 |

In the multivariate analysis we found that adults presented a time until the initial assessment significantly greater than the other age groups (p=0.048). Other variables such as sex, comorbidity diagnosis, self-medication prior to the initial assessment (NSAIDs, metamizole, corticoids), health provider and region did not significantly influence the time until initial assessment (Table 4).

**TABLE 4:**
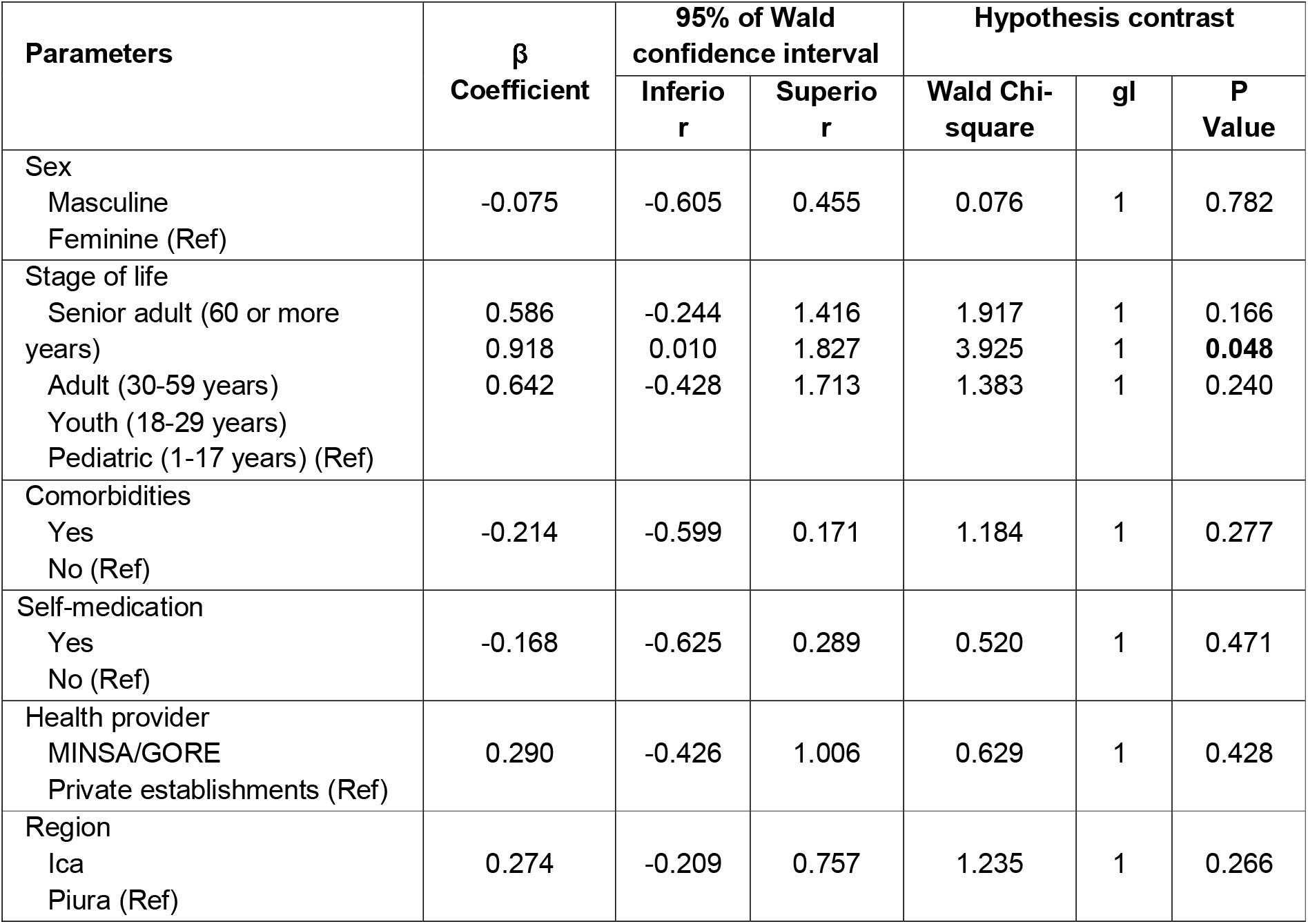
Factors that influence the elapsed time until the initial assessment of those deceased from dengue.

Regarding the time of diagnosis, we observed that the healthcare provider influenced significantly (p=0.014), observing shorter times in those seen in MINSA/GORE establishments. On the other hand, the stage of life also significantly influenced the time of diagnosis (p=0.018) observing longer times in pediatric deaths (Table 5).

**TABLE 5:** Factors that influence time of diagnosis of those deceased from dengue.

| Parameters | $\beta$<br>Coefficient | 95% of Wald<br>confidence interval | | Hypothesis contrast | | |
| --- | --- | --- | --- | --- | --- | --- |
|  |  | Inferior | Superior | Wald Chi-square | gl | p-Value |
| Sex<br>Masculine<br>Feminine (Ref) | -0.301 | -1.193 | 0.591 | 0.438 | 1 | 0.508 |
| Stage of life<br>Senior adult (60 or more years) | 0.045 | -1.424 | 1.515 | 0.004 | 1 | 0.952 |
| Youth (18-29 years) | -0.019 | -1.469 | 1.431 | 0.001 | 1 | 0.979 |
| Pediatric (1-17 years) | 2.266 | 0.389 | 4.142 | 5.602 | 1 | **0.018** |
| Adult (30-59 years) (Ref) |  |  |  |  |  |  |
| Comorbidities<br>Yes | 0.927 | -0.096 | 1.950 | 3.157 | 1 | 0.076 |
| No (Ref) |  |  |  |  |  |  |
| Health provider<br>MINSA/GORE<br>Private establishments (Ref) | -1.630 | -2.926 | -0.334 | 6.078 | 1 | **0.014** |
| Region<br>Ica<br>Piura (Ref) | -1.028 | -2.220 | 0.163 | 2.861 | 1 | 0.091 |

For the case of time of diagnosis-death, the only factor that significantly influenced was the self-medication with NSAIDs, metamizole or corticoids (p=0.041) observing shorter times in those deceased that presented self-medication prior to the initial assessment (see table 6).

**TABLE 6:** Factors that influence over time of diagnosis-death in those deceased from dengue.

| Parameter | $\beta$<br>Coefficient | 95% of Wald<br>confidence interval | | Hypothesis contrast | | |
| --- | --- | --- | --- | --- | --- | --- |
|  |  | Inferior | Superior | Wald Chi-square | gl | p-Value |
| Sex<br>Masculine<br>Feminine (Ref)) | 0.076 | -0.710 | 0.861 | 0.036 | 1 | 0.850 |
| Stage of life<br>Senior adult (60 or more years) | -0.860 | -2.305 | 0.586 | 1.359 | 1 | 0.244 |
| Adult (30-59 years) | -0.683 | -2.323 | 0.956 | 0.667 | 1 | 0.414 |
| Youth (18-29 years)<br>Pediatric (1-17 years)(Ref) | 0.049 | -1.664 | 1.762 | 0.003 | 1 | 0.955 |
| Comorbidities<br>Yes<br>No (Ref) | 0.603 | -0.179 | 1.385 | 2.281 | 1 | 0.131 |
| Self-medication<br>Yes<br>No (Ref) | -1.108 | -2.172 | -0.044 | 4.165 | 1 | **0.041** |
| Health provider<br>MINSA/GORE<br>Private establishments (Ref) | 0.358 | -0.782 | 1.499 | 0.379 | 1 | 0.538 |
| Region<br>Ica<br>Piura (Ref) | 0.538 | -0.283 | 1.360 | 1.651 | 1 | 0.199 |

## DISCUSSION

This research shows that factors exist which significantly influence the times up to the initial assessment, diagnosis and diagnosis-death of those deceased from dengue in Piura and Ica, during El Niño Costero and the Cyclone Yaku climatic-atmospheric phenomena. The adults presented a time to initial assessment significantly greater than the other age groups; the time of diagnosis was significantly less in those seen in MINSA/GORE establishments and greater in the pediatric deceased. Furthermore, the time from diagnosis to death was significantly less in those who self-medicated with NSAIDs, metamizole, or corticoids before the initial assessment. This has important implications since the FENG 2023-2024 has started, which could have similar or worse consequences if the Peruvian health system organization and capacity for resolution doesn’t improve, especially in dengue Endo epidemic areas.

The dengue epidemic in Peru during the FENC and Cyclone Yaku climatic-atmospheric phenomena, the greatest dengue epidemic in the history of Peru, demonstrated a similar pattern as COVID-19 during the pandemic. In this manner, the demand of dengue cases exceeded the health services, mainly in the north of the country^13,14,16-18^. It is possible that when no medical care was found in the health establishments, the people with dengue opted for self-medicating, which could have influenced the times and delays^19,20^.

The median of time from onset of symptoms until the initial assessment was 3 days which was slightly less than that described by Linn^21^ in a hospital in Myannmar in which the median of said time was 3 days. 20.0% of deceased presented a delay in the time until initial assessment, which is greater than that reported by Abello^22^, who found that 11.6% of severe dengue cases from an epidemiological surveillance system case series from the Philippines presented a delay until hospital admission. On the other hand, it is less than that reported by Agrawal^23^ in a teaching hospital from India where the delay reached 36.7% of severe dengue cases (including deaths). It should be emphasized that no standard exists regarding time in which a delay until initial assessment should be considered, some researchers consider it as of 5 days from the onset of symptoms and others as of 6 days. We considered a delay of 5 days due to the existence of studies that showed severe complications from the fifth day of symptomatic disease^24,25^.

The median of time until the initial assessment was 0 days (less than 1 day) which is similar to that reported by Linn^21^ (also 0 days), with the shortest time in both studies. The time until the initial assessment was significantly greater in adults which shows that adults take longer to get to health establishments than other age groups. This could be because when they are in a financially active age, they prioritize work activities and leave their health second place. Our results differed from those obtained by Wongchidwan^26^ in a hospital in Thailand, which found that the feminine sex was the only factor associated with a delay in the search for medical care (greater or equal to 5 days), however, few current studies exist that evaluate factors which influence time to the initial assessment.

The time of diagnosis was significantly less in those seen in MINSA/GORE establishments. This could be due to the experience gained by the doctors who attended in public hospitals of endemic regions, mainly in hospitals that faced important dengue epidemics that, despite their limitations in human resources, equipment, supplies, and reagents, they have organized prioritizing the diagnosis and clinical management^16,19^.

The time of diagnosis was significantly greater in the pediatric deceased. This is explained because the first days of infection, dengue presents nonspecific symptoms and signs that can be confused with those of other febrile childhood diseases that could hinder diagnosis and could lead to incorrect diagnosis and delay in treatment. This, in turn, could increase the frequency of complications, severity and mortality^27-29^.

The time of diagnosis-death was notably less than those reported by Luque^30^ in the deceased from dengue during the dengue epidemic that occurred in Piura, Peru in 2017, when Peru was found to be under the effects of FENC. Considering that Luque reported that hte median of time from arrival at the establishment to death was 4.1 days and that said time in our study was composed by the sum of time of diagnosis (median of 0 days) and the time of diagnosis-death (median of 1 day), during the climatic-atmospheric phenomena El Niño Costero and the Cyclone Yaku in 2023 a notably shortening of time of survival of deceased, meaning, that patients died much faster in 2023 than in 2017.

The time of diagnosis-death was significantly less in those who self-medicated with NSAIDs, metamizole or corticoids prior to the initial assessment. This implies that once the diagnosis was made, those who self-medicated with these drugs died faster than those who did not self-medicate. A possible explanation of this phenomena is found in the growing tendency of self-medicating in Peru which could negatively affect the clinical course of diseases such as dengue when anti-inflammatories and fever-reducers are used without a medical prescription which could trigger or worsen the hemorrhagic manifestations and lead to hypovolemic shock ^30-32^.

Almost all deceased presented warning signs during the initial assessment, despite that, dengue was not suspected in an important fraction of patients. The study carried out by Luque^30^ about clinical and epidemiological characteristics of dengue deaths in Piura during the FENC of 2017 found that the percentage of other diagnosis during admission in the first health establishment was notably less (16.7% versus 37.5%). A possible explanation for not suspecting dengue on behalf of clinicians could be the presence of nonconventional clinical patterns simulating other febrile diseases^29^, another possible explanation was the frequent association of dengue and advanced age and comorbidities which represent an added challenge for the diagnosis^33,34^. It is also possible that during the FENC 2023/Cyclone Yaku, the presentation of dengue cases had an unusual behavior, with a modified clinical presentation modified by circulating viral and bacterial coinfections (whose incidence increased with FENC and Cyclone Yaku) of which prior reports also exist^35-37^.

Our article has limitations that need to be discussed. The most important entails the small number of deaths considered, however, our study included more than half of deaths occurred in Piura and Ica during the effects of FENC and the Cyclone Yaku. Furthermore, we should take into account that under normal conditions, the number of deaths from dengue is low. Another limitation is that a non-probabilistic sample of consecutive cases was carried out which could affect the sample representation. Finally, since this study was performed from secondary sources, the possibility exists that there is some level of underreporting, which was partially controlled with the use of diverse sources of information.

In conclusion, adults presented a significantly greater time until initial assessment, the time of diagnosis was less in those seen in MINSA/GORE establishments, and greater in pediatrics. Furthermore, the time from diagnosis until death was significantly less in those who self-medicated before the initial assessment. An important fraction of deceased presented comorbidities and an unconventional clinical pattern during admission with a frequent lack of suspicion of dengue during the initial assessment.

## Data Availability

Data may be made available by contacting the corresponding author.

